# HPV16 and HPV18 type-specific APOBEC3 and integration profiles in different diagnostic categories of cervical samples

**DOI:** 10.1101/2020.11.25.20238477

**Authors:** Sonja Lagström, Alexander Hesselberg Løvestad, Sinan Uğur Umu, Ole Herman Ambur, Mari Nygård, Trine B. Rounge, Irene Kraus Christiansen

**Author notes:** Equal contribution (Trine B. Rounge, Irene Kraus Christiansen). Corresponding authors E-mail addresses (TBR), (IKC).

## Abstract

Human papillomavirus 16 and 18 are the most predominant types in cervical cancer. Only a small fraction of HPV infections progress to cancer, indicating that genomic factors, such as minor nucleotide variation caused by APOBEC3 and chromosomal integration, contribute to the carcinogenesis.

We analysed minor nucleotide variants (MNVs) and integration in HPV16 and HPV18 positive cervical samples with different morphology. Samples were sequenced using an HPV whole genome sequencing protocol TaME-seq. A total of 80 HPV16 and 51 HPV18 positive cervical cell samples passed the sequencing depth criteria of 300× reads, showing the following distribution: non-progressive disease (HPV16 n=21, HPV18 n=12); cervical intraepithelial neoplasia (CIN) grade 2 (HPV16 n=27, HPV18 n=9); CIN3/adenocarcinoma in situ (AIS) (HPV16 n=27, HPV18 n=30); cervical cancer (HPV16 n=5).

Similar rates of MNVs in HPV16 and HPV18 samples were observed for most viral genes but for HPV16, the non-coding region (NCR) showed a trend towards increasing variation with increasing lesion severity. APOBEC3 signatures were observed in HPV16 lesions, while similar mutation patterns were not detected for HPV18. The proportion of samples with integration was 13% for HPV16 and 59% for HPV18 positive samples, with a noticeable portion located within or close to cancer-related genes.

## 1. Introduction

A persistent infection with one of the carcinogenic HPV genotypes is accepted as a necessary cause of cervical cancer development [1]. Of the 12 carcinogenic types [2], HPV16 and HPV18 are associated with about 70% of all cervical cancers [3]. HPV16 is predominantly associated with squamous cell carcinomas (SCC), while HPV18 is more often detected in adenocarcinomas [3], suggesting that these HPV types differ in their target cell specificity [4]. Nevertheless, only a small fraction of HPV infections will persist and progress to cancer [5], indicating that additional factors and genomic events are necessary for the HPV-induced carcinogenic process.

The 7.9 kb double stranded HPV DNA genome consists of early region (E1, E2, E4-7) genes, late region (L1, L2) genes and an upstream regulatory region (URR) [6]. To date, more than 200 HPV genotypes have been identified, based on at least 10% difference within the conserved L1 gene sequence [7]. HPV types harbouring minor genetic variation are grouped into lineages (1–10% whole genome nucleotide difference) and sublineages (0.5-1.0% difference) [8]. HPV evolve slowly partly since the HPV genome replication is dependent on host cell high-fidelity polymerases [9]. However, recent studies have revealed variability below the level of HPV sublineages, which may indicate intra-host viral diversification and evolution [10-12].

Viral genetic instability is caused by various mutagenic processes [13]. One of the mechanisms suggested to cause minor nucleotide variants (MNVs) and impact HPV mutational drift involves the anti-viral host-defence enzyme apolipoprotein B mRNA-editing enzyme, catalytic polypeptide-like 3 (APOBEC3) protein [14]. APOBEC3 proteins are cytidine deaminases causing deoxycytidine (C) to deoxythymidine (T) mutations during viral genome synthesis. The mutations can lead to defects in viral genome replication necessary for the viral life cycle [15]. APOBEC3 mutational signatures have been found in HPV genomes in cervical pre-cancerous and cancer samples [10, 16, 17], and has recently been associated with viral clearance [18]. Findings of hypovariability of the E7 gene suggest negative selection opposite of APOBEC3-related editing and an essential gene conservation for progression to cancer [19, 20].

HPV integration into the host genome is regarded as a driving event in cervical carcinogenesis and is observed in >80% of HPV-induced cancers [21]. Integrations causing disruption or complete deletion of the E1 or E2 gene result in constitutive expression of the viral E6 and E7 oncogenes [22], leading to inactivation of cell cycle checkpoints and genomic instability [23]. Integration may also lead to disruption of host genes, such as oncogenes or tumour-suppressor genes, modified expression of adjacent genes, as well as other genomic alterations, which may promote HPV-induced carcinogenesis [24-26]. In high-grade lesions and cancers, integrations in certain chromosomal loci, including loci 3q28, 8q24.21 and 13q22.1, have been reported more often than in other loci [27], suggesting selective growth advantages for cells with site-specific integrations in e.g. important regulatory genes. Increasing integration frequencies have been reported upon comparison of cervical precancerous and cancer lesions [28, 29].

Recently, we developed a novel next-generation sequencing (NGS) strategy TaME-seq for simultaneous analysis of HPV genomic variability and chromosomal integration [30]. Employing the TaME-seq method, we have explored HPV16 and HPV18 genomic variability and integration in HPV positive cervical samples with different morphologies. Differences in HPV variability between the diagnostic categories may shed light on intra-host viral genome dynamics and evolution processes in cervical carcinogenesis. In addition, integration analysis will contribute to a better understanding of this event during HPV-induced carcinogenesis.

## 2. Material and methods

### 2.1. Sample selection

Cervical cell samples were collected from women attending the cervical cancer screening program in Norway between January 2005 and April 2008. Samples were collected in ThinPrep PreservCyt solution (Hologic, Marlborough, MA) and pelleted before storage at −80 °C. The samples were stored in a research biobank at Akershus University Hospital, consisting of both the cell material and extracted DNA. Recruitment criteria and HPV detection and genotyping have been described previously [31, 32]. Cytology samples were previously analysed for HPV using the Amplicor HPV DNA test (Roche Diagnostics, Switzerland) followed by genotyping by Linear Array (Roche Diagnostics, Switzerland) and PreTect HPV-Proofer (PreTect AS, Norway).

In this study, primarily DNA was used for downstream analyses; for some samples, DNA extraction had to be performed from the cell material. DNA extraction was performed using the automated NucliSENS easyMag platform (BioMerieux Inc., France) with off-board lysis. All samples in the biobank that were positive for HPV16 and/or HPV18, alone or together with other HPV types, by one or both of the genotyping methods were included in the study, with the exception of HPV16 CIN3 samples for which a random selection of 50 samples were included. In total, 157 HPV16 positive cytology samples and 75 HPV18 positive samples were subjected to sequencing (**Table 1**). All samples were allocated to mutually exclusive categories based on the HPV type and the diagnostic categories of non-progressive disease, histologically confirmed cervical intraepithelial neoplasia (CIN) grade 2 (CIN2), CIN3/adenocarcinoma *in situ* (AIS) and cancer. Non-progressive disease category included samples with normal cytology that also had normal cytology the preceding two years and with no previous history of treatment for cervical neoplasia (HPV16 n=24, HPV18 n=3), and samples with atypical squamous cells of undetermined significance (ASC-US) or low-grade squamous intraepithelial lesions (LSIL) with no follow-up diagnosis within four years subsequent to the diagnosis (HPV16 n=31, HPV18 n=13). For the CIN2, CIN3/AIS and cancer categories, sequencing was performed on cell samples taken at the time of conisation; cytological examination of these samples was not performed. The cancer category included SCC (n=4) and adenocarcinoma (n=1) samples.

**Table 1.**
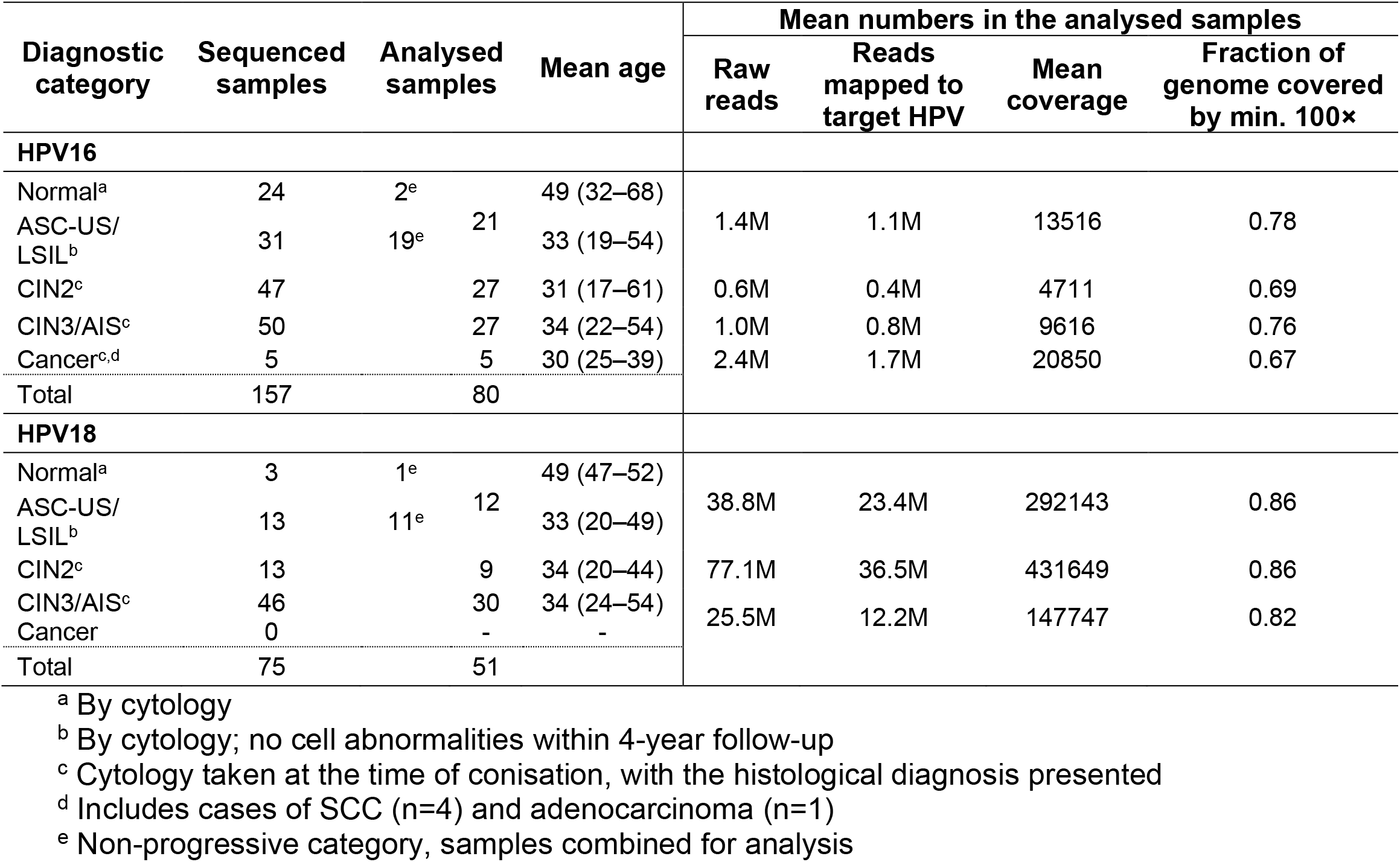
Number of samples, and mean mappings statistics in each HPV16 and HPV18 diagnostic category.

### 2.2. Library preparation and sequencing

Library preparation was performed using the TaME-seq method as described previously [30]. In brief, samples were subjected to tagmentation using Nextera DNA library prep kit (Illumina, Inc., San Diego, CA), following target enrichment performed by multiplex PCR using HPV primers and a combination of i7 index primers [33] and i5 index primers from the Nextera index kit (Illumina, Inc., San Diego, CA). Sequencing was performed on the HiSeq2500 platform with 125 bp paired-end reads.

### 2.3. Sequence alignment

Data was analysed by an in-house bioinformatics pipeline as described previously [30]. Reads were mapped to human genome (GRCh38/hg38) using HISAT2 (v2.1.0) [34]. HPV16 and HPV18 reference genomes were obtained from the PaVE database (https://pave.niaid.nih.gov). Mapping statistics and sequencing coverage were calculated using the Pysam package [35] with an in-house Python (v3.5.4) script. Downstream analysis was performed using an in-house R (v3.5.1) script. Samples with a mean sequencing depth of <300× were excluded from the further analysis.

### 2.4. Sequence variation analysis

Mapped nucleotide counts over the HPV reference genomes and average mapping quality values for each nucleotide were retrieved from the HISAT sequence alignment. Variant calling was performed using an in-house R (v3.5.1) script. Nucleotides seen ≤2 times in each position and nucleotides with mean Phred quality score of <20 were filtered out. Both F and R nucleotide counts from the same sample, obtained independently from separate amplification reactions, were combined and variant allele frequencies were calculated for each genomic position. If variants called from the two separate reactions were discordant, the highest covered variant was used. Genomic positions covered with <100× were filtered out. Variants were called if the variant frequency was >1%.

The ratio of non-synonymous to synonymous substitutions (dN/dS) was calculated to indicate potential positive (new variants favoured) or negative (new variants eliminated) selection affecting protein-coding genes. For mutational signature analysis, all nucleotide substitutions were classified into six base substitutions, C>A, C>G, C>T, T>A, T>C, and T>G, and further into 96 trinucleotide substitution types, including information on the bases immediately 5’ and 3’ of the mutated base. Analysis was performed using an in-house R (v3.5.1) script.

### 2.5. Detection of chromosomal integration

Integration site detection was performed as described previously [30]. In brief, a two-step analysis strategy was employed to identify read pairs spanning integration sites. First, read pairs with one read mapped to HPV and the other to the human chromosome were identified using HISAT2. Second, unmapped reads were re-mapped using the LAST (v876) aligner (options -M -C2) [36] to increase detections of the above mentioned read pairs. Reads sharing the same start and end coordinates were considered as potential PCR duplicates and were excluded. Selected integration sites were confirmed by PCR amplification and Sanger sequencing on the ABI^®^ 3130xl/3100 Genetic Analyzer 16-Capillary Array (Thermo Fisher Scientific Inc., Waltham, MA) using BigDye™ Terminator v1.1 cycle sequencing kit (Thermo Fisher Scientific Inc., Waltham, MA). Samples with a mean depth of >1000× and <85% of the genome covered by minimum 100× were manually inspected using IGV (v2.3.09) to detect HPV genomic deletions.

### 2.6. Functional annotation of genes within or close to integration sites

Nearest gene, with a transcription start site within 100 kb from the integration site, was identified using Ensembl. Gene2function (http://www.gene2function.org) and Genecards (https://www.genecards.org) were used to annotate the molecular function and disease phenotype of each gene. SNP associations in the GWAS Catalog [37] were retrieved from Genecards. Genes involved in cell cycle regulation, cell proliferation, apoptosis, tumour suppressor mechanisms, cancer-related pathways, or genes interacting with these pathways, or genes with direct cancer-related SNP associations were termed as cancer-related genes. The integration sites were manually inspected using Geneious Prime (v.2019.0.4) to investigate whether the integration site was located in exons, introns or UTRs. Information regarding regulatory elements, including promoters, promoter flanking regions, enhancers and CTCF-binding sites, was retrieved from Ensembl regulatory build [38]. Integration sites in retained introns, ncRNA and antisense RNA were reported if they had a transcript support level of 1 or 2.

### 2.7. Statistical analysis

Statistical analyses were done in R (v3.5.1). The Kruskal-Wallis test was used to examine differences in numbers and frequencies of MNVs and integrations between the groups. A p-value of <0.05 was considered statistically significant.

### 2.8. Ethical considerations

This study was approved by the Regional Committee for Medical and Health Research Ethics, Oslo, Norway (REK 2017/447). Written informed consent has been obtained from all study participants.

## 3. Results

### 3.1. Characteristics and sequencing statistics

This study included 232 HPV16 and HPV18 positive cytology cell samples which were categorised according to cytology or histology diagnosis. A total of 80 HPV16 positive samples and 51 HPV18 positive samples, allocated to diagnostic categories of non-progressive disease, CIN2, CIN3/AIS and cancer, passed the strict sequencing depth criteria necessary for further analyses of minor nucleotide variation and integration. In total, 1.05 billion read pairs were analysed. The mean sequencing coverage in the categories ranged from 4711 (CIN2) to 20850 (cancer) for HPV16 positive samples and from 147747 (CIN3/AIS) to 431649 (CIN2) for HPV18 positive samples. On average, the samples had 77.7% of the genome covered with a minimum depth of 100× (**Table 1**).

### 3.2. Minor nucleotide variation profiles similar for HPV16 and HPV18

Overall, the number of MNVs was similar in HPV16 and HPV18 positive samples, and between the diagnostic categories. In total, 3747 MNVs were found in all 131 samples. In HPV16 positive samples, the mean number of variants found in the non-progressive category was 37 per sample, 30 in the CIN2 category, 28 in the CIN3/AIS category, and 25 in the cancer category. Corresponding numbers for HPV18 positive samples were 24, 21, and 27 for the non-progressive, CIN2 and CIN3/AIS categories, respectively (**Figure 1A**). HPV16 positive samples had mean MNV frequencies of 2.9% for non-progressive, 3.2% for CIN2, 3.6% for CIN3/AIS and 3.7% for cancer categories. For HPV18 positive samples, the mean MNV frequencies were 3.1% for non-progressive, 2.6% for CIN2 and 5.0% for CIN3/AIS categories (**Figure 1B**). Statistical analysis was performed; the mean numbers and MNV frequencies were not statistically different between the HPV types or the diagnostic groups within an HPV type.

**Figure 1.**
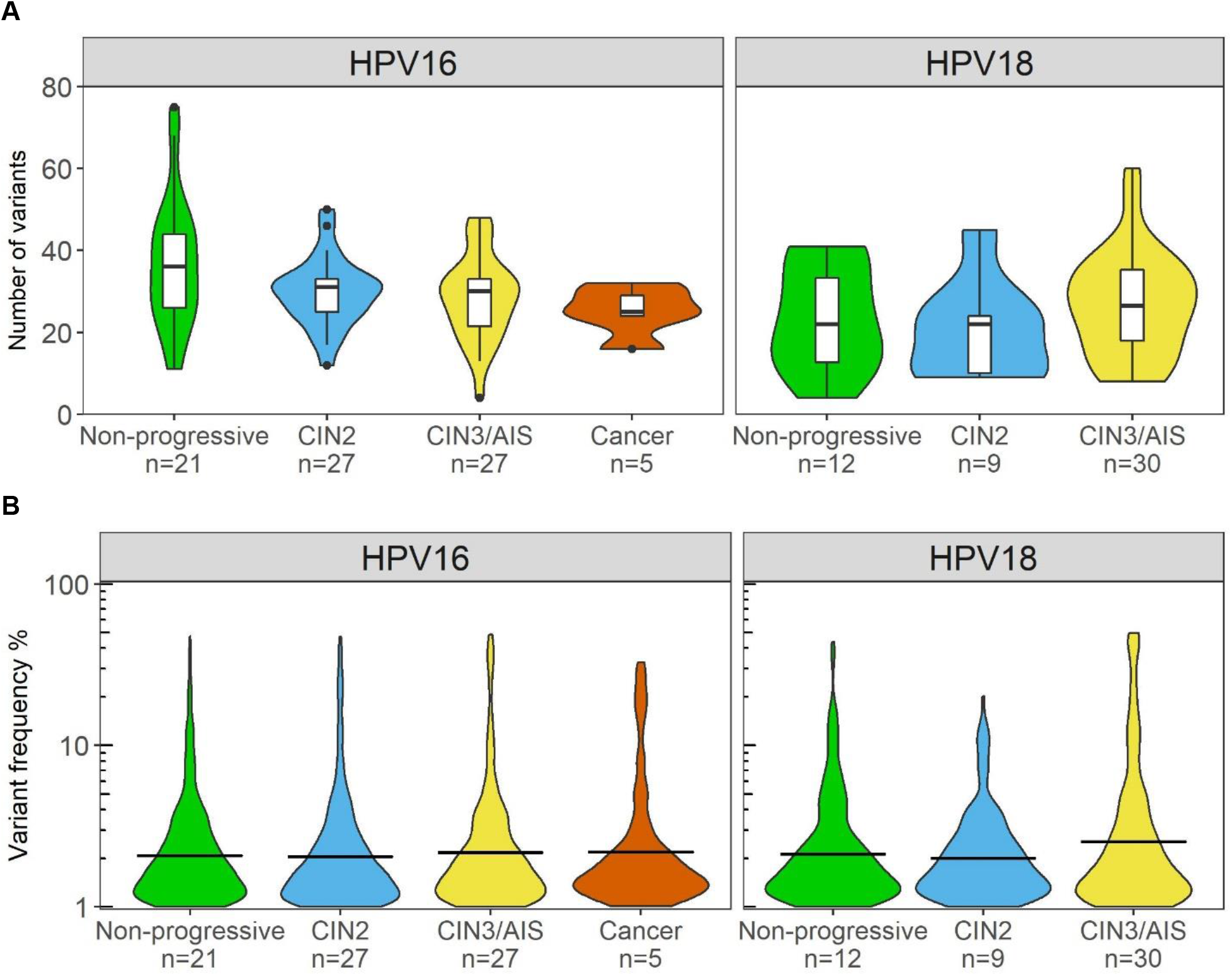
Number of variants and variant frequencies in HPV16 and HPV18 positive samples. A) Number of variants presented as violin plots across the different diagnostic categories. Boxplots are added to show the mean number and distribution of variants. B) Variant frequencies (%) of detected minor variants shown as violin plots across the different diagnostic categories. The vertical bar indicates the mean variant frequency.

### 3.3. Different level of variation in HPV16 and HPV18 genes

HPV variants occurred throughout all HPV genes (**Figure 2A**). Notably, of HPV16 genomic regions, the short non-coding region (NCR) between E5 and L2 showed increased minor nucleotide variation. The nucleotide variation showed clear trend to be higher in NCR than in any other genomic region in diagnostic categories CIN2, CIN3/AIS and cancer but this could not been tested statically due to the low number of observations in each category. A higher degree of variation was also observed in the HPV18 E4 gene. The dN/dS patterns for HPV16 showed mostly nonsynonymous variants (dN/dS > 1), while a considerable part of HPV18 genes had equal amounts of nonsynonymous and synonymous variants (dN/dS ≈ 1) (**Figure 2B**). Strikingly, several HPV16 genes showed signs of positive selection, i.e. a preference for non-synonymous mutations (dN) over synonymous mutations (dS). HPV16 E6 had the most pronounced dN/dS ratio of 6. In contrast, the E7 gene in the same samples had a dN/dS ratio of 0.4, indicating neutral or negative selection. In HPV18 positive samples, most nonsynonymous substitutions were observed in the E2 gene, with a dN/dS ratio of > 2 in all diagnostic categories. For the other HPV18 genes, the dN/dS ratio was close to 1 across diagnostic categories.

**Figure 2.**
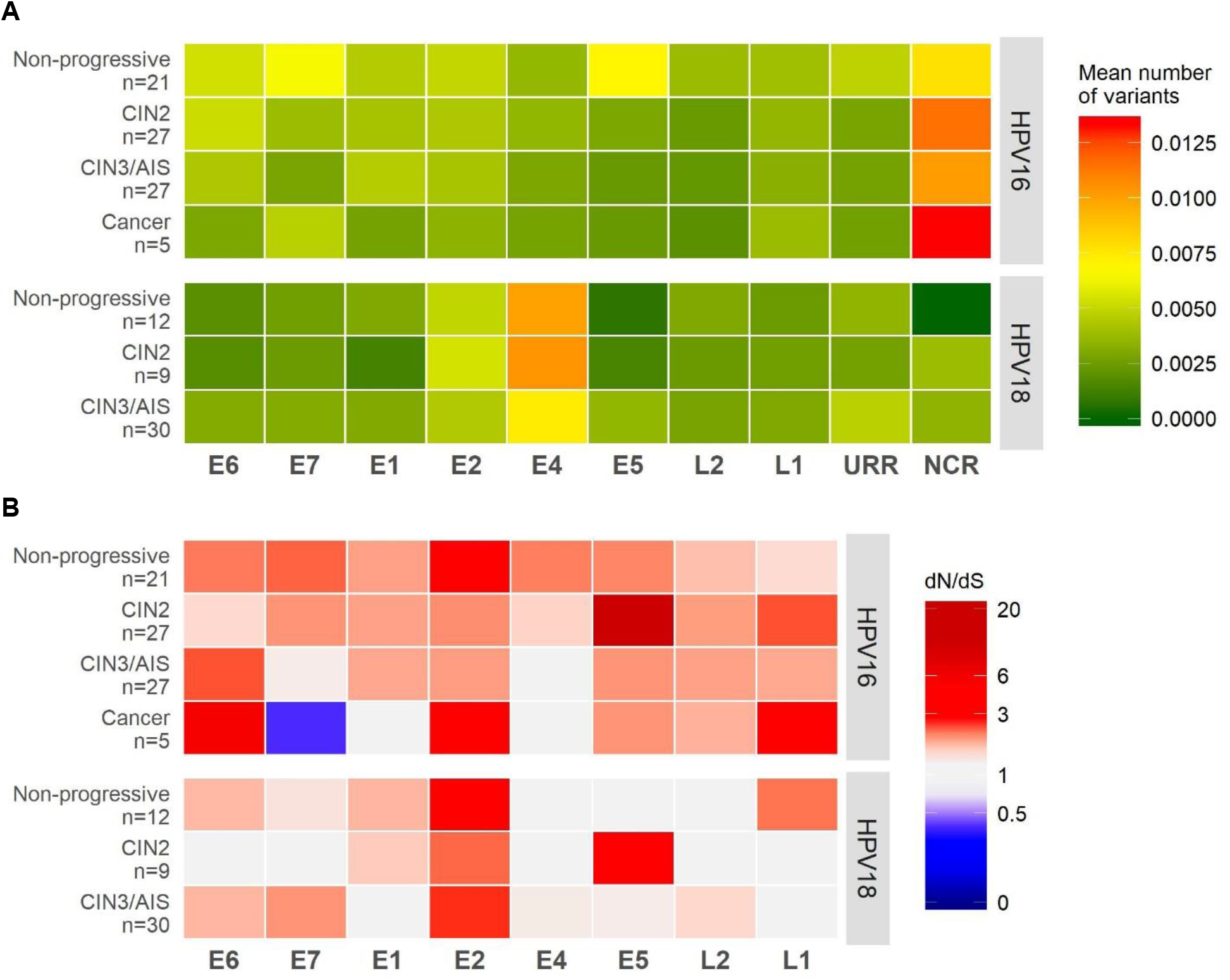
Number of variants, nonsynonymous and synonymous variations in the different HPV genes. A) Heat map with green-yellow-red gradient color-coding representing mean number of variants per sample in HPV16 and HPV18 genomic regions. Number of variants is normalised by the gene length and stratified by the diagnostic category. B) Heat map with blue-white-red gradient color-coding representing the ratio of non-synonymous to synonymous substitutions (dN/dS) in HPV16 and HPV18 genomic regions across the different diagnostic categories.

### 3.4. APOBEC3-related mutational signatures identified in non-progressive and CIN2 samples

Among nucleotide substitutions, predominantly C>T and T>C substitutions were observed across all diagnostic categories (**Supplementary Figure S1**). The APOBEC3-related C>T substitutions were compared between the different categories and HPV types (**Figure 3**). C>T substitutions in the trinucleotide context TCW (W is A or T), a preferred target sequence for the APOBEC3 proteins [39] and a more stringent motif than TCN (N is any nucleotide [40], was the most prevalent mutational signature type in HPV16 non-progressive samples and to a slightly less extent in HPV16 CIN2 samples. HPV16 CIN3/AIS and cancer samples did not show any preferred signature patterns. Interestingly, HPV18 samples showed different C>T trinucleotide substitution patterns compared to HPV16 samples. In all HPV18 diagnostic categories, C>T substitutions in the trinucleotide context ACA was predominantly observed, while C>T substitutions in the trinucleotide context GCA was the second most prevalent in non-progressive and CIN2 samples.

**Figure 3.**
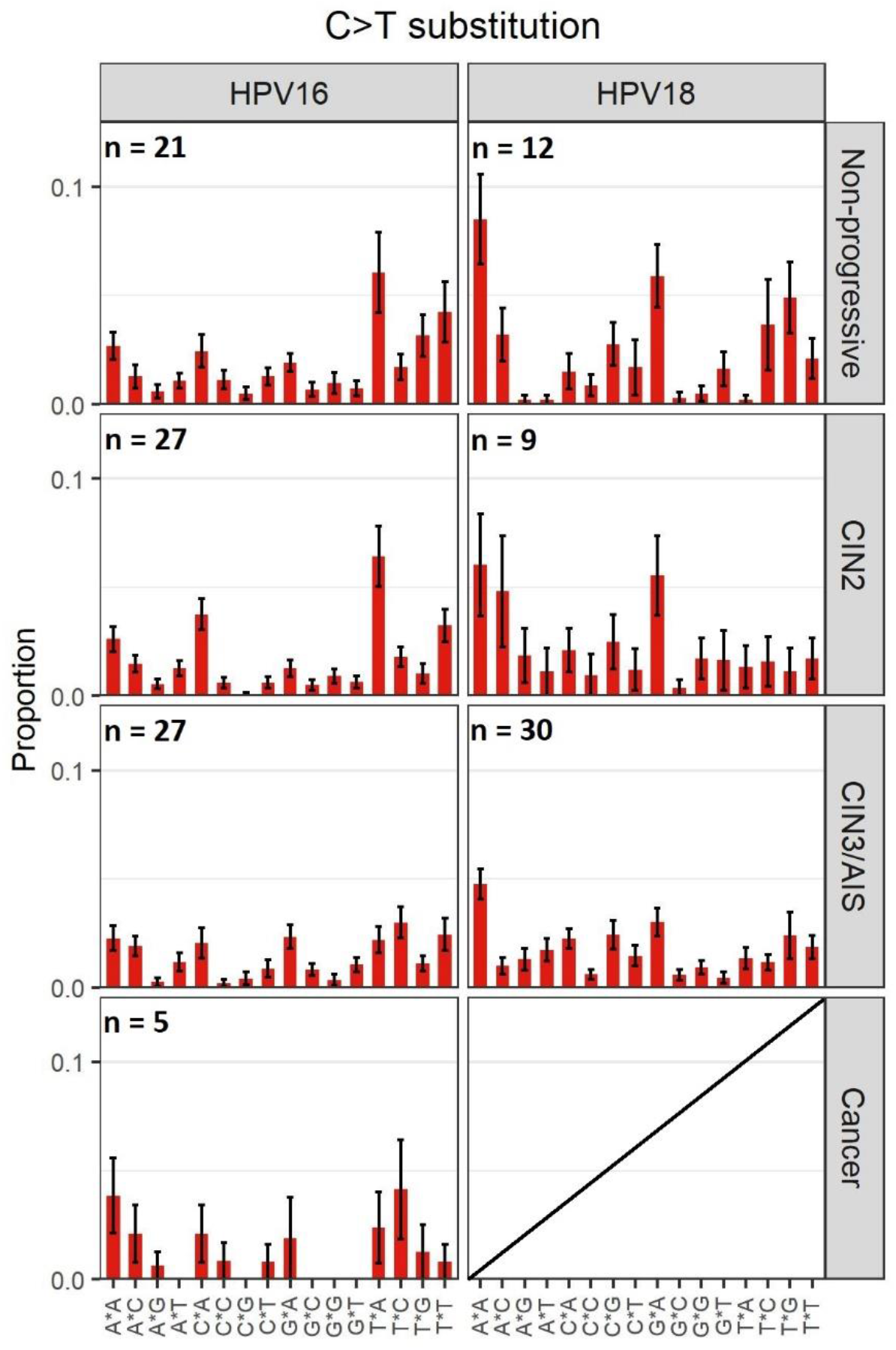
C>T mutational signatures in HPV16 and HPV18 positive samples. The mean proportion of 16 trinucleotide substitution types is shown across the different diagnostic categories. Error bars represent the standard error of the mean.

### 3.5. Higher HPV integration frequencies in HPV18 than in HPV16 positive samples

The proportion of samples with integration was 13% (10/80) for HPV16 and 59% (30/51) for HPV18 positive samples (**Table 2**). The integration frequency was higher in all HPV18 positive diagnostic categories compared to the HPV16 categories. Of the HPV16 positive samples, HPV integration was detected in 4%, 7% and 60% in CIN2, CIN3/AIS and cancer samples, respectively. Corresponding numbers in HPV18 samples were 78% and 53% for CIN2 and CIN3/AIS categories, respectively. The total number of integration sites found in each diagnostic category was in general higher for HPV18 positive samples, ranging from 22 (CIN2) to 60 (CIN3/AIS), while for HPV16 samples, a total of 17 integration sites were identified (**Table 2**).

**Table 2.** Number of HPV16 and HPV18 positive samples with integration, stratified by the diagnostic categories.

| Diagnostic category | Number of samples<br>with integration<br>(Frequency %) | Total number<br>of integration<br>sites |
| --- | --- | --- |
| **HPV16** |  |  |
| Non-progressive (n=21) | 4 (19%) | 5 |
| CIN2 (n=27) | 1 (4%) | 2 |
| CIN3/AIS (n=27) | 2 (7%) | 3 |
| Cancer (n=5) | 3 (60%) | 7 |
| Total (n=80) | 10 (13%) | 17 |
| **HPV18** |  |  |
| Non-progressive (n=12) | 7 (58%) | 24 |
| CIN2 (n=9) | 7 (78%) | 22 |
| CIN3/AIS (n=30) | 16 (53%) | 60 |
| Total (n=51) | 30 (59%) | 106 |

In Figure 4A, the difference between HPV16 and HPV18 positive samples in terms of number of integration sites is illustrated, stratified by diagnostic category. Combined for all diagnostic groups, HPV18 samples had significantly more integration sites than HPV16 samples (p-value < 0.001). The mean numbers of integration sites per HPV18 positive sample were 3.4, 3.1 and 3.8 for the non-progressive, CIN2 and CIN3/AIS categories, respectively. The mean numbers of integration sites per HPV16 positive sample with observed integration, were 1.3, 2, 1.5 and 2.3 for the non-progressive, CIN2, CIN3/AIS and cancer categories, respectively (**Figure 4A**). In total, six HPV16 positive samples and 18 HPV18 positive samples had more than one integration site observed (**Supplementary Table S1**).

**Figure 4.**
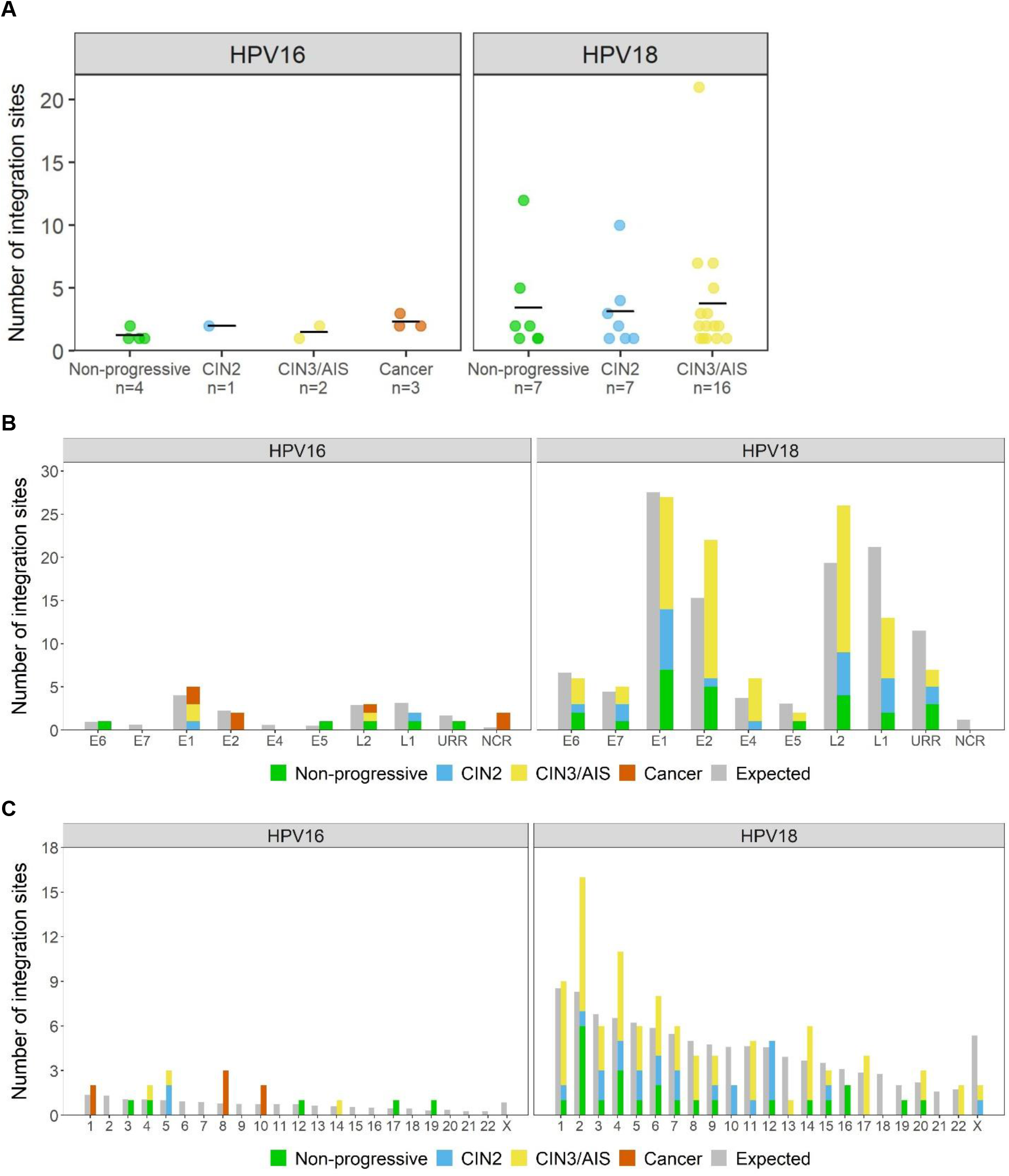
Integration sites in HPV16 and HPV18 positive samples. A) Number of integration sites in samples with observed integration. Each spot in the plot indicates one sample, and total number of samples with integration is specified for each diagnostic category. Vertical lines indicate the mean number of integration sites. B) Integration sites in HPV genes. C) Integration sites in human chromosomes and expected number of breakpoints with regard to randomness is presented.

The validation rates of integration sites using Sanger sequencing (good quality chromatograms produced) was 44% (7/16 samples) (**Supplementary Table S1, Supplementary Table S2**). A PCR product or a smear was identified on agarose gel but no clean chromatogram was seen in additional 44% (7/16) of the reactions (**Supplementary Figure S2**). Two integration sites, one in HPV16 and one in HPV18 positive sample, both in the non-progressive category, could not be confirmed (**Supplementary Table S1**).

### 3.6. Integration sites and deletions in the HPV genome

For HPV16, integrations in the viral genome were detected in all genes except E4 and E7. Remarkably, NCR between the E5 and L2 genes, harboured two integration sites in one cancer sample (**Figure 4B, Supplementary Table S1**). In the HPV18 positive samples, integration sites were located in all HPV genomic regions except NCR. Expected number of integrations in each gene relative to gene lengths was estimated with regard to randomness by dividing the total number of integration sites within a HPV type by the length of the gene. Based on this, integration was more frequently observed in E1 and NCR in HPV16 samples and in E2, E4 and L2 in HPV18 samples, while L1 and URR were less prone to integration (**Figure 4B**). For HPV16 and HPV18 combined, integration sites were located in E1 or E2 in 38%, 38%, 48%, and 57% of all the integration sites in non-progressive, CIN2, CIN3/AIS, and cancer categories, respectively (**Supplementary Figure S3**). All cancer samples had at least one integration site in E1 or E2 (**Supplementary Table S1**).

HPV genomic regions covered with very few or no sequencing reads were considered as deletions according to previous validations [30]. Such deletions were observed in six samples; one HPV16 positive sample (cancer) and five HPV18 positive samples (**Supplementary Figure S4**). For these samples, human sequences were detected flanking the deleted regions, indicating chromosomal integration. In all six samples, the genomic deletion encompassed the region between E1/E2 and L2. The deletions were either partial, suggesting the presence of both episomal and integrated HPV DNA, or complete with no reads detected for the deleted region.

### 3.7. Integration sites in the human genome

In HPV16 positive samples, integration sites (n=17) were distributed on 10 chromosomes; for the cancer samples, all integration sites (n=7) were located on chromosomes 1, 8 or 10 (**Figure 4C**). Interestingly, the integration sites on chromosome 8 were located in the *PVT1* oncogene, in the chromosomal locus 8q24.21 (**Supplementary Table S1**), previously being defined as an HPV integration hotspot [27]. For the HPV18 positive samples, integration sites (n=106) were found in all chromosomes except chromosomes 18 and 21 (**Figure 4C**). Most HPV18 integration sites were observed on chromosomes 2 and 4. In HPV18 samples, 36% (4/11) of the integration sites on chromosome 4 were located in the previously defined hotspot locus 4q13.3 [27], all from samples diagnosed with CIN2 or CIN3/AIS.

Due to a low frequency of integration events in HPV16 positive samples, HPV16 and HPV18 samples were combined for reporting HPV integrations affecting different human genetic elements. The frequency of integration sites located in human genes ranged from 50 to 71%, with the highest frequency observed in cancer samples (**Figure 5A**). Integration sites were detected in or close to cancer-related genes (**Supplementary Table S3**) in 100% (7/7) of cancer samples (n=3), in 65% (41/63) of CIN3/AIS samples (n=18), in 38% (9/24) of CIN2 samples (n=8), and in 34% (10/29) in non-progressive samples (n=11) (**Figure 5B**). In individual samples, the highest numbers of integration sites located in or near cancer-related genes was 13/21 in CIN3/AIS, 3/10 in CIN2, and 5/12 in non-progressive samples, all being HPV18 positive (**Supplementary Figure S5**).

**Figure 5.**
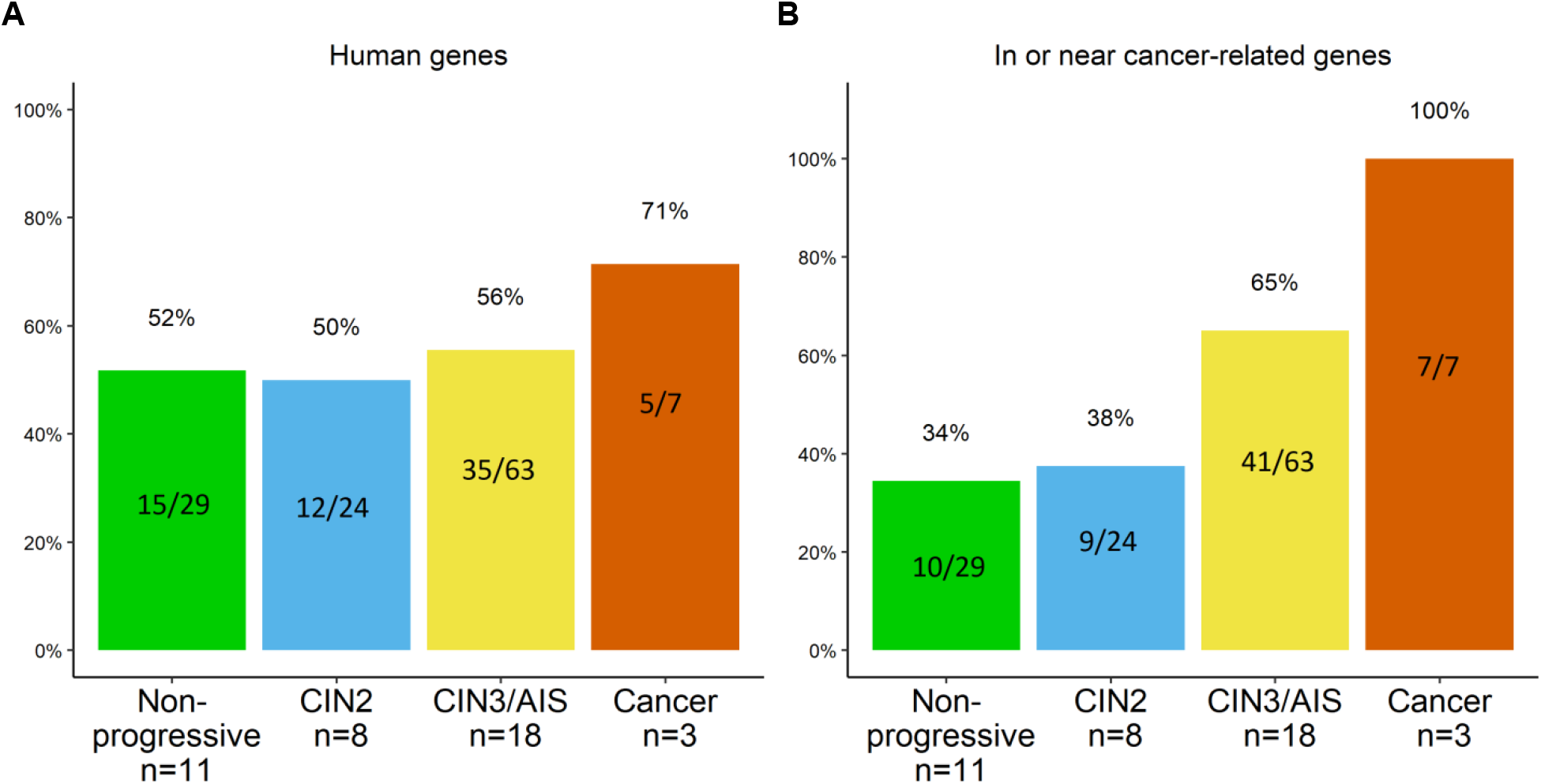
The frequency of integration sites combined for HPV16 and HPV18 A) in human genes, and B) in or near human cancer-related genes. Number of integration sites is indicated inside the bars and total number of samples with integration (n) is specified for each diagnostic category.

Integration located in exons, introns, regulatory regions, retained introns, non-coding RNA (ncRNA), antisense RNA and untranslated regions (UTRs) varied between the diagnostic groups (**Supplementary Table S1**). Integration frequency in exons and regulatory regions decreased with lesion severity, while the integration frequency in introns, retained introns and ncRNA increased with lesion severity. Antisense and UTR showed only few integrations in certain diagnostic groups (**Supplementary Figure S6**).

## 4. Discussion

This study compares HPV16- and HPV18-associated genomic events in samples showing no clinical progression, CIN2, CIN3/AIS and cervical cancer samples. We find that the genomic events differ dependent of lesion severity but more importantly that these are strikingly different between HPV16 and HPV18 positive samples. To our knowledge, we are the first to show that with increasing lesion severity variation in NCR increases and report, in line with other studies [10, 18], decreases in APOBEC3-related nucleotide substitutions in HPV16 positive samples. As previously reported [21, 41], HPV18 samples show higher integration frequencies compared to HPV16, while we found an increase in integration frequencies in or in close proximity to cancer-related genes with increasing lesion severity.

In this study, the number and frequency of MNVs was similar between HPV genotypes and morphological categories. Recent HPV deep sequencing studies, exploring HPV genomic variation with various PCR-based NGS approaches and different variant calling thresholds, show slightly divergent numbers of MNVs [10, 18, 30]. We found a total of 3747 MNVs in the 131 samples, being in line with studies reporting a high number of HPV variation at the population level [20, 42, 43], within infected hosts [10, 11, 30]. A recent study on HPV16 genome stability analysed possible HPV16 sublineage co-infections and observed 20–38 variants in each sample [44], corresponding to the mean numbers of MNVs in this study. The variation was reported not to be due to co-infections but interpretation of the nucleotide variation source was not further elaborated [44].

For HPV16, a higher degree of variation in the NCR was observed in the categories CIN2, CIN3/AIS and cancer. This was not seen for HPV18 positive samples. Recent studies document high degrees of variation in HPV16 NCR, but without any biological interpretation.[10, 19] The NCR in HPV16 has been characterised to portray a weak promoter activity specific to L2 mRNA expression [45]. Repeat sequences of varying length in NCR have been reported [46] and the NCR has been shown to harbour miRNA binding sites [47]. The loss of miRNA binding sites due to nucleotide variation in NCR was suggested to serve as a novel mechanism to sustain L2 expression, and thereby justify the potential role of L2 in HPV-induced carcinogenesis [47]. However, an opposite finding has also been reported, showing more variation in NCR in clearing than in persistent HPV16 infections [42]. Interestingly, we also observed integration sites in NCR in one cancer sample, which may indicate involvement of NCR in HPV16-induced carcinogenesis. To our knowledge, this is the first study to report integration in NCR.

Ratio of nonsynonymous to synonymous variants (dN/dS) is used as indicator of positive or negative selection occurring over generations [13]. This ratio may indicate non-random occurrence of minor nucleotide variability. In this study, the observed nucleotide variations in the HPV16 and HPV18 genes were biased toward nonsynonymous substitutions, being in line with previous results showing a high ratio of non-synonymous nucleotide variation [10]. Only HPV16 E7 had a dN/dS ratio of <1, indicating negative selection and conservation of function. Interestingly, two recent studies reported similar results on strict conservation of the HPV16 E7 gene at the population level [19, 20]. A potential source of the substitutions may be the APOBEC3 enzyme creating C>T substitutions through its cytidine deaminase activity [15]. APOBEC3-related mutations have previously been reported in cervical cancer lesions [10, 17, 18]. Our finding of APOBEC3-related signatures in the HPV16 positive non-progressive samples indicates that this mechanism is active also in an early stage of infection. The relative amount of variants related to APOBEC3 may at a more severe stage of disease disappear, due to an increase in non-APOBEC3 mutations caused by e.g. hampered DNA repair mechanisms in an increasingly cancerous environment [48]. This study was the first to characterise mutational patterns in HPV18 samples, showing mutation patterns in the trinucleotide context RCA (R is A or G), a target motif for the activation-induced cytidine deaminase (AID) that is a member of the APOBEC protein family [49].

HPV-induced carcinogenesis is a multi-step process that may be facilitated through the disruption of host genes and genomic instability by viral integration [24-26]. A high number of integrations in a sample may in itself be a sign of genomic instability, which may further accelerate such events. In our dataset, multiple integration sites were observed in 24 samples, with the maximum of 21 integration sites in one HPV18 sample in the CIN3/AIS category, possibly promoting a higher degree of chromosomal instability. Our results showed a higher number of integration events in HPV18 positive samples compared to HPV16 positive samples, being consistent with previous observations [21, 41]. Genomic instability as a consequence of multiple integrations, is further strengthened by finding integrations in the E1 and E2 genes, which might result in overexpression of the viral oncogenes E6 and E7. Previous studies using NGS methodology for HPV integration analysis report disruptions mainly in E1 and E2 genes [50, 51]. In addition, we found genomic deletions in one HPV16 positive cancer sample and in five HPV18 positive samples of all categories. In all of these, the genomic deletion always led to partial or complete loss of E1, E2 and L2. Similar results showing HPV genomic deletions have been reported in cervical carcinomas [52] and HPV positive oropharyngeal squamous cell carcinomas [53].

Due to the low frequency of integration events in HPV16 positive samples, HPV16 and HPV18 integrations were combined for the analysis of integrations in or close to cancer-related genes. We observed an overall increase in the proportion of integration sites within or close to cancer-related genes with increasing lesion severity. All integrations in the cancer samples occurred within or near the cancer-related genes *PVT1, WAC* and *miR-205*. The *PVT1* oncogene, a long non-coding RNA gene, has been associated with multiple cancers including cervical cancer [54]. The *PVT1* gene is located in the chromosomal locus 8q24.21, which is one of the regions previously reported to contain integration sites in cervical carcinomas more often than other loci [27]. Transcription of *PVT1* is regulated by the key tumour suppressor protein p53 and PVT1 is implicated in regulating the *MYC* oncogene [55]. The WAC protein regulates the cell-cycle checkpoint activation in response to DNA damage and is a positive regulator of mTOR, which functions as a key player in the regulation of cell growth and metabolism [56]. The miRNA miR-205 has been implicated in many cancers and it targets genes involved in DNA repair, cell cycle control and cancer-related pathways [57]. In the CIN2 and CIN3/AIS categories, 38% and 65% of the integration sites were observed in or close to cancer-related genes, respectively. Interestingly, integration sites in or close to cancer-related genes were also observed in the non-progressive disease category. Whether this might represent one of several components for risk stratification remains to be determined. Our results, together with a recent study [58], have shown that viral integrations may also occur in other genetic elements that are involved in regulation of gene expression, such as ncRNA and UTRs.

NGS protocols with comprehensive analyses of whole HPV genomes, their variability and integrations, enable greater understanding of the role of genomic events during cancer development. By comparatively analysing genomic events, we get a broader picture of the dynamic changes in the HPV genome during malignant cell transformation. HPV16 and HPV18 are to a certain degree associated with different types of invasive cervical cancers [3, 4] and may utilise different molecular mechanisms to induce carcinogenesis. Firstly, HPV18 is suggested to cause more genomic instability [4, 41] and HPV18 lesions are more likely to progress from CIN3 to cancer than HPV16 positive lesions [4]. Furthermore, previously reported results show different DNA methylation patterns [59] and mechanistic signatures of integrations [52] for HPV16 and HPV18, which strengthens the hypothesis of different underlying mechanisms for HPV16- and HPV18-induced cervical carcinogenesis.

Despite the large sample number in total, the sample size in certain diagnostic categories was low, limiting us from performing statistical analyses and drawing conclusions from the given part of the dataset. Some samples, mainly in the non-progressive category, had low sequencing coverage for the HPV genome. This is most likely explained by low viral load, which was not measured in the samples. Low viral load has previously been observed to affect the sequencing yield [12]. Two integration sites in non-progressive samples were not confirmed by Sanger sequencing. This may be explained by unsuccessful PCR primer design, suboptimal PCR conditions, or may reflect repeated integrations or other genomic structures affecting the PCR reaction. Still, since the NGS data showed clear results, both integration sites were included in the analysis.

## 5. Conclusions

To summarise, we have in this study analysed HPV minor nucleotide variation, chromosomal integration and genomic deletions in cervical cell samples with different morphology by utilising the TaME-seq protocol [30]. The results show a high number of low-frequency variation, distinct variation patterns and integration frequencies, providing initial insight into dissimilar genomic alterations between HPV16 and HPV18, possibly reflecting differences in the mechanisms of cell transformation induced by the two genotypes. In addition, the study adds to the growing evidence of within-host HPV genomic variability. Cancer registry data with information on future cervical disease or longitudinal studies including patient outcome, preferably with a larger sample size for all diagnostic categories, are needed for further interpretation of different HPV whole genome MNV signatures and to validate the role and importance of viral integrations.

## Data Availability

The data presented in this article are not readily available because of the principles and conditions set out in the General Data Protection Regulation (GDPR), with additional national legal basis as per the Regulations on population-based health surveys and ethical approval from the Norwegian Regional Committee for Medical and Health Research Ethics (REC). Requests to access the data should be directed to the corresponding authors.

## Abbreviations

AID: activation-induced cytidine deaminase
AIS: adenocarcinoma *in situ*
ASC-US: atypical squamous cells of undetermined significance
CIN: cervical intraepithelial neoplasia
dN/dS: ratio of non-synonymous to synonymous substitutions
HPV: human papillomavirus
LSIL: low-grade squamous intraepithelial lesion
MNV: minor nucleotide variant
NCR: non-coding region
ncRNA: non-coding RNA
NGS: next-generation sequencing
SCC: squamous cell carcinoma
URR: upstream regulatory region
UTR: untranslated region

## Acknowledgements

We thank Mona Hansen for DNA extraction, Hanne Kristiansen for sequencing library preparation, Karin Helmersen for Sanger sequencing, and Marcin W. Wojewodzic for his help with gene annotation of the chromosomal integration sites.

## Authors’ contributions

SL designed and performed the experiments, analysed the results and drafted the manuscript text. AL analysed the results and contributed to drafting the manuscript. SUU contributed to the data analysis. OHA contributed to the study design and result interpretation. MN contributed to the clinical interpretation of the results. TBR contributed to the study design, data analysis and result interpretation. IKC managed the sample material, contributed to the study design and result interpretation. All authors contributed to writing and approved the final version of the manuscript.

