## Supplementary figures for "HPV16 and HPV18 type-specific APOBEC3 and integration profiles in different diagnostic categories of cervical samples"

**A**

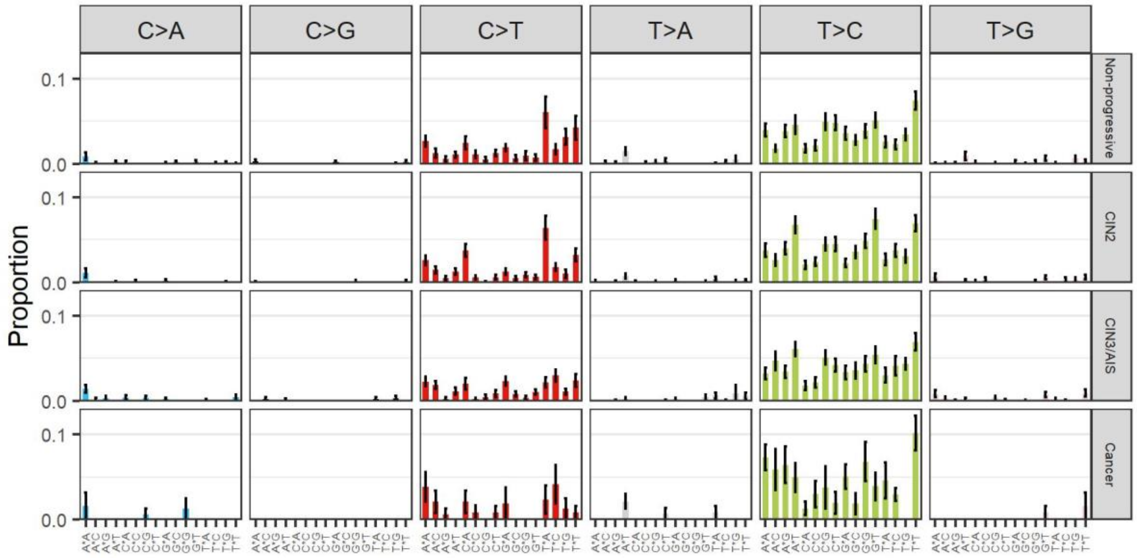


**B**

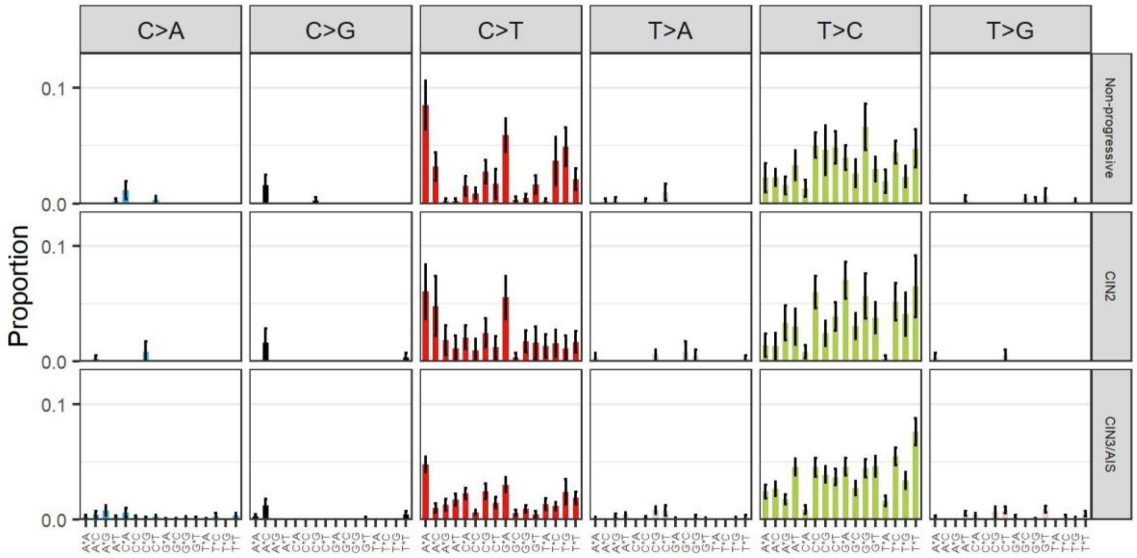


**Supplementary Figure S1.** Mutational signatures in A) HPV 16, and B) in HPV18 positive samples. All substitutions are classified into six base substitutions and the mean proportion of 96 trinucleotide substitution types is shown across the different diagnostic categories. Error bars represent the standard error of the mean.

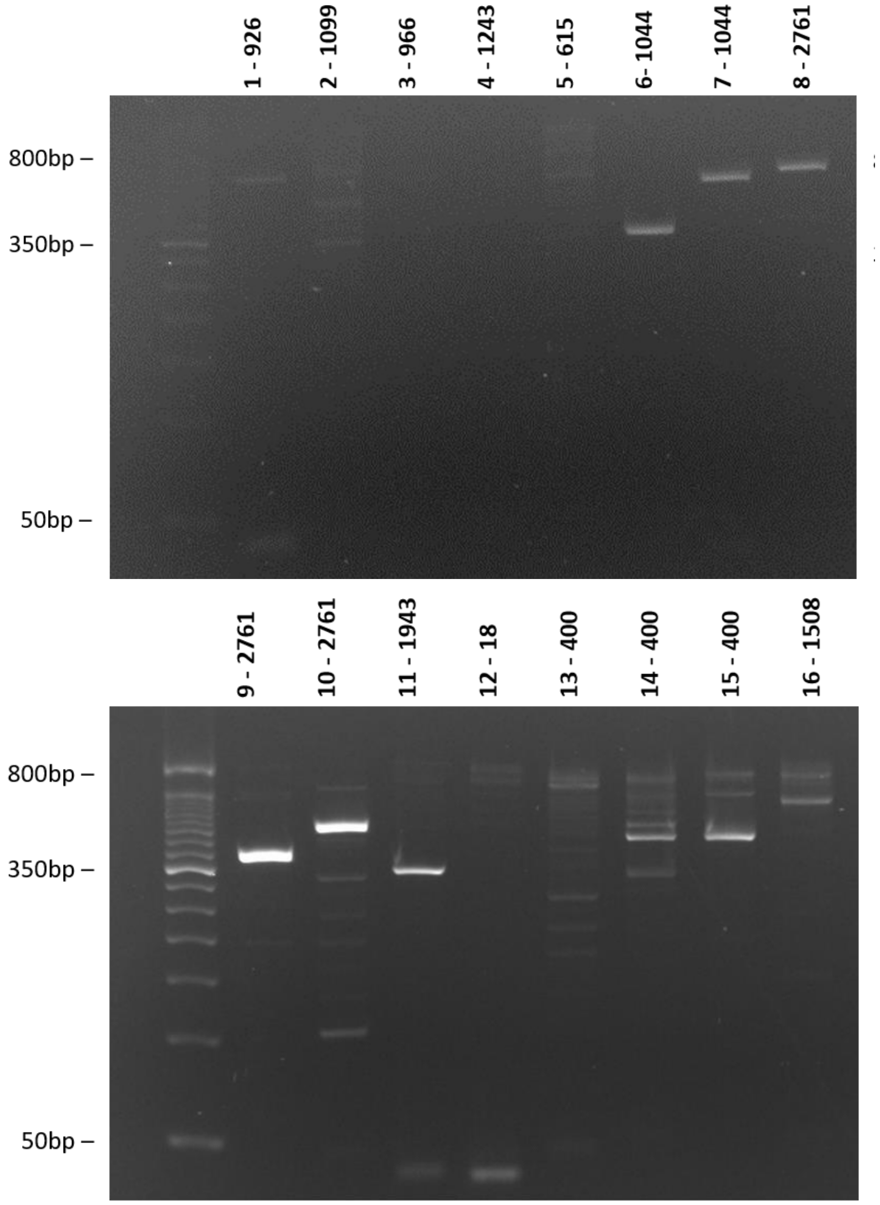


**Supplementary Figure S2.** Integration validation PCR products on agarose gel.

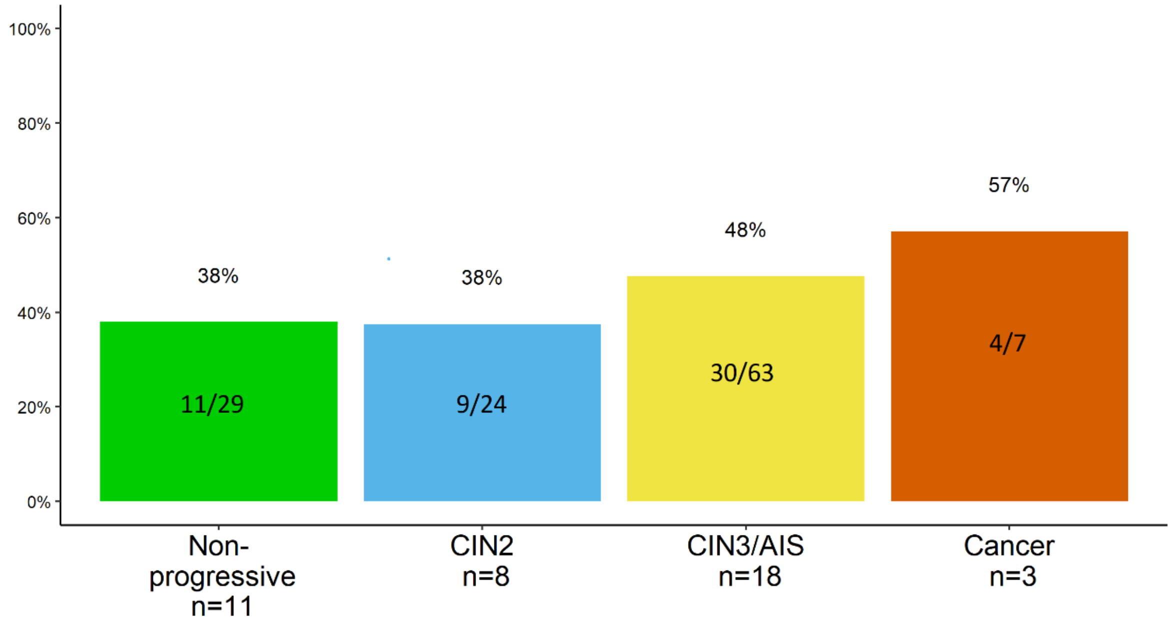


**Supplementary Figure S3.** The frequency of integration sites in E1 or E2 stratified by the diagnostic categories. Number of integration sites is indicated inside the bars and total number of samples with integrations (n) is specified for each diagnostic category.

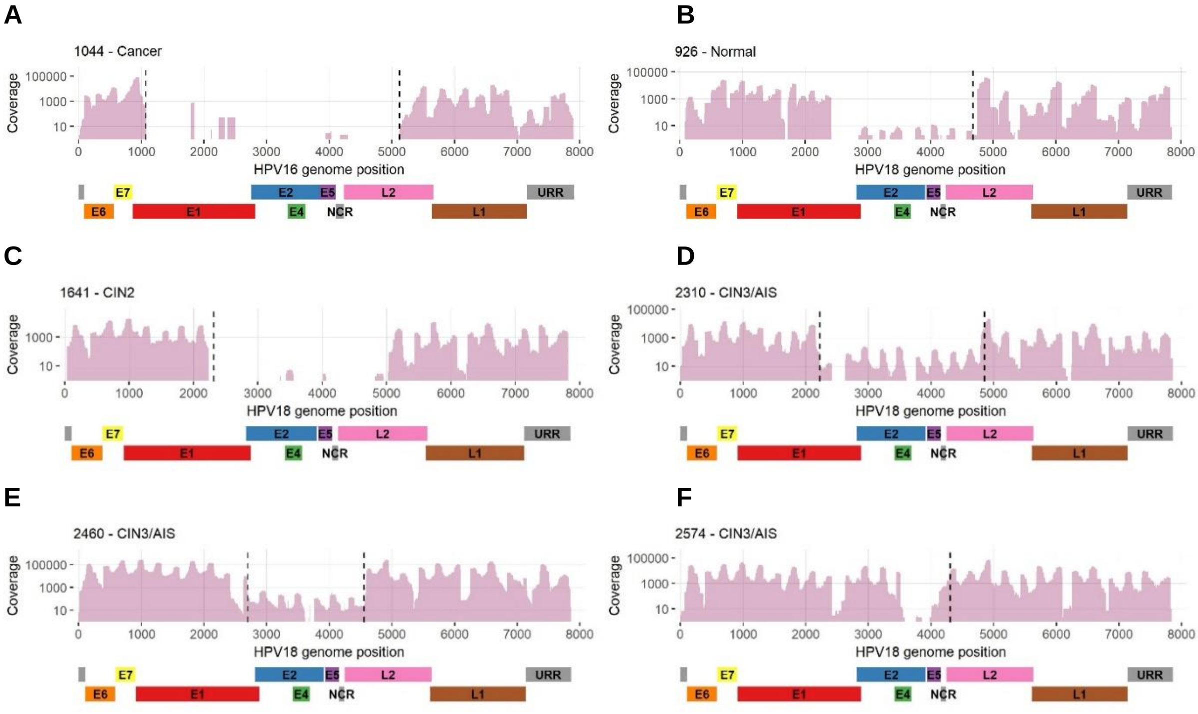


**Supplementary Figure S4.** Genomic deletions in HPV16 (A) and HPV18 positive samples (B-F). The respective regions were deleted completely (no sequencing coverage) or partially (low sequencing coverage). Dashed vertical lines indicate the integration sites detected with the integration analysis.

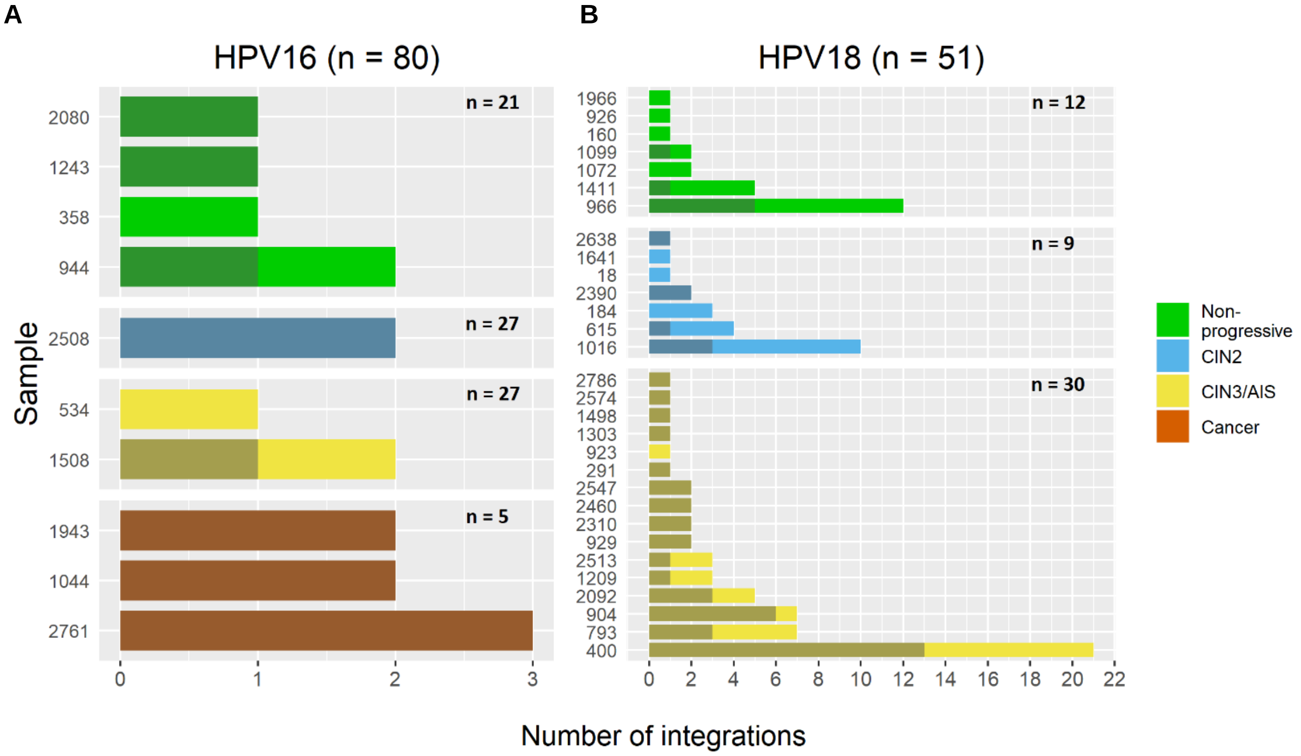


**Supplementary Figure S5.** Number of integration sites in A) HPV16 positive and B) HPV18 positive samples. Darker area in each bar indicates integration sites within or near cancer-related genes. In the cancer category, all sites occurred in or close to cancer-related genes. Total number of samples is indicated in the plot for each diagnostic category.

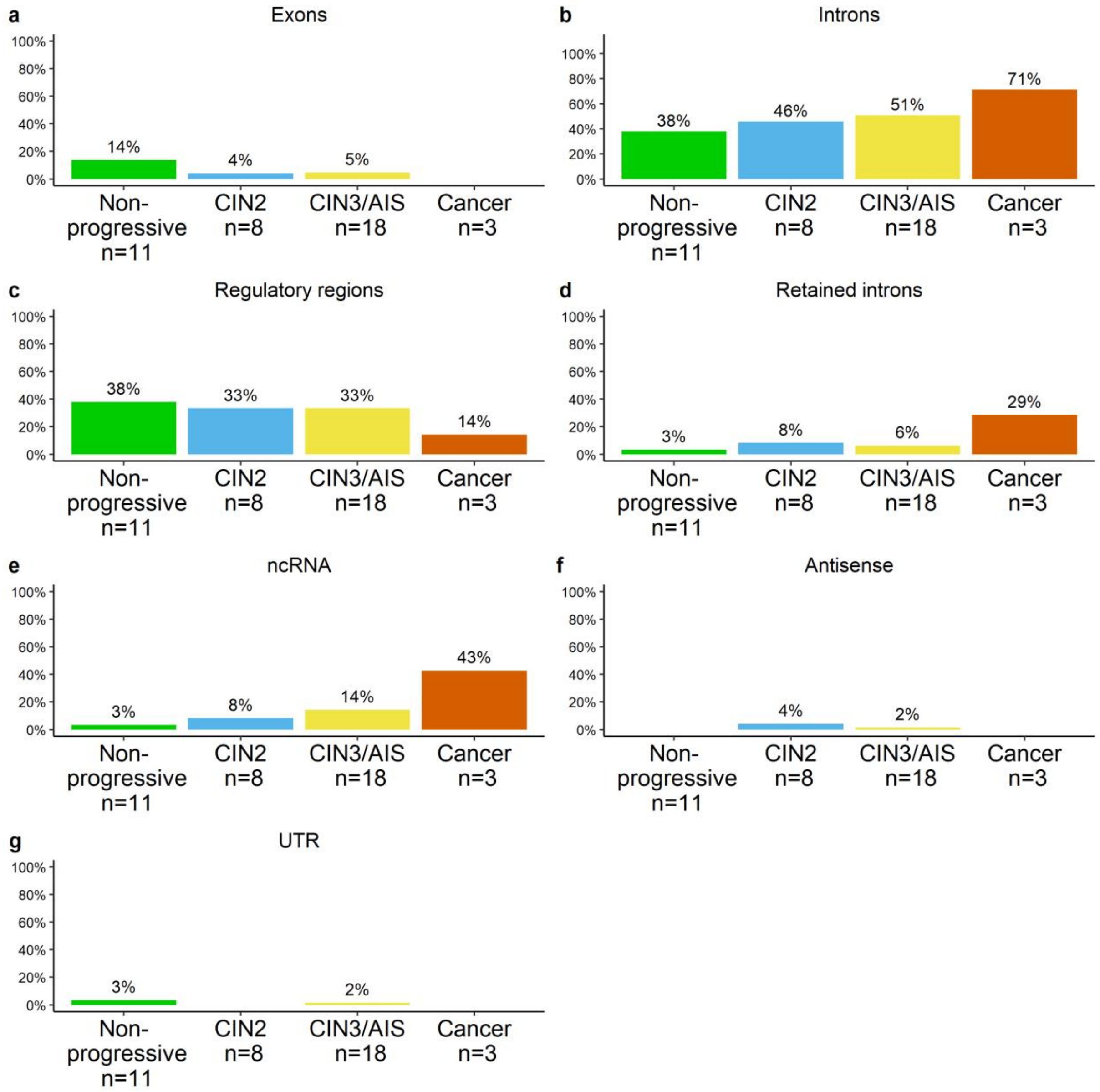


**Supplementary Figure S6.** The frequency of integration sites in A) exons, B) introns, C) regulatory regions, D) retained introns, E) ncRNA, F) antisense RNA, G) UTR. Number of samples with integration (n) are specified for each diagnostic category.
